# SARS-CoV-2-positive patients display considerable differences in proteome diversity in urine, nasopharyngeal, gargle solution and bronchoalveolar lavage fluid samples

**DOI:** 10.1101/2022.01.08.22268611

**Authors:** Javan Okendo, Clarisse Musanabaganwa, Peter Mwangi, Martin Nyaga, Harris Onywera

## Abstract

**Background:** Proteome profile changes post-severe acute respiratory syndrome coronavirus 2 (post-SARS-CoV-2) infection in different body sites of humans remains an active scientific investigation whose solutions stand a chance of providing more information on what constitutes SARS-CoV-2 pathogenesis. While proteomics has been used to understand SARS-CoV-2 pathogenesis, there are limited data about the status of proteome profile in different human body sites infected by the sarscov2 virus. To bridge the gap, our study aims to profile the proteins secreted in urine, bronchoalveolar lavage fluid (BALF), gargle solution, and nasopharyngeal samples and assess the proteome differences in these body samples collected from SARS-CoV-2-positive patients.

**Materials and methods:** We downloaded publicly available proteomic data from (https://www.ebi.ac.uk/pride/). The data we downloaded had the following identifiers: i) PXD019423, n=3 from Charles Tanford Protein Center in Germany. ii) PXD018970, n=15 from Beijing Proteome Research Centre, China. iii)PXD022085, n=5 from Huazhong University of Science and Technology, China, and iv) PXD022889, n=18 from Department of Laboratory Medicine and Pathology, Mayo Clinic, Rochester, MN 55905 USA. MaxQuant was used for the peptide spectral matching using humans, and SARS-CoV-2 was downloaded from the UniProt database (access date 13^th^ October 2021).

**Results:** The individuals infected with SARS-CoV-2 viruses displayed a different proteome diversity from the different body sites we investigated. Overall, we identified 1809 proteins across the four different sample types we compared. Urine and BALF samples had significantly more abundant SARS-CoV-2 proteins than the other body sites we compared. Urine samples had 257(33.7%) unique proteins, followed by nasopharyngeal with 250(32.8%) unique proteins. Garage solution and BALF had 38(5%) and 73(9.6%) unique proteins.

**Conclusions:** Urine, gargle solution, nasopharyngeal, and bronchoalveolar lavage fluid samples have different protein diversity in individuals infected with SARS-CoV-2. Moreover, our data demonstrated that a given body site is characterized by a unique set of proteins in SARS-CoV-2 seropositive individuals.

## 1. Introduction

Severe acute respiratory syndrome coronavirus 2 (SARS-CoV-2) emerged in Wuhan, China, in December 2019 and spread rapidly worldwide (1). The SARS-CoV-2 is an enveloped RNA virus with a positive-sense, single-stranded RNA genome of approximately 30 kb (*Coronaviridae* Study Group of the International Committee on Taxonomy of Viruses, 2020). Coronavirus disease 2019 (COVID-19) is the clinical syndrome associated with SARS-CoV-2 and is characterized by respiratory or gastrointestinal viral symptoms. COVID-19 may result in clinical features such as cardiovascular, neurological, thrombosis, and renal failure (2). Approximately 269 million COVID-19 cases with over 5.3 million deaths were reported globally on 13-12-2021 (https://CoVid19.who.int/). Globally, the exact impact of SARS-CoV-2 infection is unknown even though the number confirmed is still in the upward trajectory. The number of cases seems to vary geographically. As of 13^th^ December 2021, the approximate number of cases per region were as follows America: 98 million, Europe: 91 million, Southeast Asia: 44 million, Eastern Mediterranean: 16 million, Western pacific: 10 million, and Africa: 6.5 million (https://CoVid19.who.int/).

COVID-19 spread from person to person through direct contact or encountering infected surfaces. When SARS-CoV-2 is inhaled, it enters the human host cells via angiotensin-converting enzyme 2 (ACE2) receptors (3). Once the virus enters the human cells, it starts replicating, leading to population expansion within the cells (3). While in the cells, it induces the local immune cells to start producing cytokines and chemokines, resulting in the attraction of other immune cells in the lung, which causes excessive tissue damage (4). A growing body of evidence indicates that the SARS-CoV-2 virus is not confined in the human lungs. Still, it also affects the other body organs, such as the kidney, where it causes acute kidney injury (AKI) (5). In other individuals infected with SARS-CoV-2, neurological, cardiovascular, and intestinal malfunctions have also been reported (2).

Proteomics has played a fundamental role in the surveillance of SARS-CoV-2 spread globally, drug target identification, vaccine designs, and the development of rapid diagnostic kits used in health facilities (6). Proteomics has enabled the development of methods to detect SARS-CoV-2 infections to complement the genomics assays (7). Proteomic profiling aims to identify the most regulated proteins following SARS-CoV-2 infection to identify the potential biomarkers. Still, it can also be used to understand the host-SARS-CoV-2 interactions, protein-protein interactions, post-translational modifications, proteome expression patterns, and the cellular localization of the proteins (6,8). To date, functional and differential proteomics has enabled the generation of an enormous amount of information, leading to the identification and characterization of SARS-CoV-2 infection and the regulated pathways (9,10). Different proteomic biospecimen has increased our understanding of the SARS-CoV-2 virus dynamics in our population. Ihling and associates used the proteome obtained from the gargle solution to develop the mass spectrometry identification method for SARS-CoV-2 identifications (7). In predicting SARS-CoV-2 clinical outcome, urine has been used, and the motivation could be due to ease of collection, which makes it attractive for proteomic analysis (9).

On the other hand, Li and associates used urine to profile individuals with the SARS-CoV-2 infection (11). Samples collected from the nasopharyngeal site, the point of SARS-CoV-2 entry, have been integral in characterizing the host response following SARS-CoV-2 infection (8). Bronchoalveolar lavage fluid (BALF) has provided answers to poorly understood questions of the SARS-CoV-2 pathogenesis at the site of infection, the human lungs (10). Using BALF, Zheng identified pathways involved in oxidative stress and the immunological responses as the main enriched pathways from the BALF proteome (10). These studies have used biospecimen collected from different body sites, with each focusing on a specific biospecimen.

To the best of our knowledge, no documented proteomics study has attempted to understand the proteome profile of the human nasopharyngeal, bronchoalveolar space, urine, and the gargle solutions from individuals with SARS-CoV-2 infection. Therefore, we hypothesized that the proteome profile of the infected body sites is different because these tissues are made up of different cell types. In this study, we sort to gain more insight into the proteomic profiles of human urine, BALF, gargle solutions, and nasopharyngeal proteome and assess how the proteome profile compares when the SARS-CoV-2 virus colonizes different body sites.

## 2. Materials and methods

This study analyzed publicly available data downloaded from the Protein Identification Database, PRIDE (https://www.ebi.ac.uk/pride/) repository. The data used in this analysis had the following identifiers: i) PXD019423 (gargle solution, n=3) (7), ii) PXD018970 (Urine, n=15) (11), iii) PXD022085 (BALF, n=5) (10), and iv) PXD022889 (nasopharynx, n=18) (8). We acknowledge that the samples used in this study were processed in different laboratories worldwide, and the sample preparation protocols have been reported elsewhere in the respective publications (7–10).

## 3. Bioinformatics data analysis

Raw data files were processed with MaxQuant version 1.6.10.43 (12) for protein and peptide identification using the Andromeda search engine and the combined Uniprot proteome for *Homo sapiens* (Proteome ID: UP000005640, 78120 entries, and SARS-CoV-2 Proteome ID: UP000464024, entries 17 both accessed on 13/10/2021). MaxQuant default parameter settings were used for the MS/MS database search, with carbamidomethylation of cysteine residues and acetylation of protein N-termini selected as fixed modification and oxidation of methionine as variable modification. The peptide spectral matches (PSMs) were filtered at a 1 % false discovery rate (FDR), and the precursor mass tolerance was set at 20 ppm. Trypsin/P was selected as protease, label-free quantitation (LFQ) was enabled. The samples from the different studies were processed together to ensure cross-normalization, making the proteome comparison more accurate. Before the downstream analysis, reverse hits and common contaminants were removed from the data set.

We did further data processing, where we used the Bioconductor package ‘Differential Enrichment analysis of Proteomics data’ version 1.2.0 (13) to do the proteomic differential analysis by comparing the body sites under investigation. The protein groups identified in 70% of patients by at least two unique peptides were retained for analysis. We used “MinProb” for imputation with a q-value cut-off of 0.01. For unsupervised clustering, principal component analysis was performed on the data after imputation. Proteins differentially expressed after the challenge were identified using the limma function, including Benjamin Hochberg multiple testing corrections. Proteins were considered differentially expressed if they survived a log2(x) fold change of 2 (as indicated) and an adjusted p-value of 0.05. Volcano Plots were visualized using the ‘enhancedVolcanoplot’ package in the Bioconductor package.

## 4. Results

This study hypothesized that the other body sites respond differently to SARS-CoV-2 infection, and the proteome profiles differ. This is because each body site is made up of unique cell types. Thus, the proteins collected from gargle solution, nasopharyngeal, bronchoalveolar lavage fluid, and urine samples were analyzed to assess the host’s proteome profile from different body sites.

### Proteome samples clustered according to body site

The recent study using multi-omics approaches such as proteomics, transcriptomes phosphoproteome, and ubiquitinome demonstrated that SARS-CoV-2 infections cause perturbations of the host upon infection at different omics levels (14). Following SARS-CoV-2 infections in human hosts, it has been demonstrated that it affects different body sites such as epithelium layers (15), kidneys (5), enterocytes (16), and lung injuries (17). Thus, we wondered if the proteome profile from different body sites has the same proteome profile or differences in protein composition post-SARS-CoV-2 infection in humans. Our study investigated the proteome profile of the urine, bronchoalveolar lavage fluid, gargle solution, and Nasopharynx from individuals with confirmed SARS-CoV-2 infection, which were deposited on the PRIDE repository (18). We hypothesized that the host proteome profile in the different body sites is the same. The study reveals that the proteomes secreted in the different body sites have different protein profiles and compositions [Figure 1]. Overall,1809 proteins were detected across the four different sample types we compared after removing potential contaminants, reverse proteins, and the proteins identified only by sites, and the one hit wonders. The findings show that the urine samples clustered closely together following principal component analysis (PCA) suggesting that the urine proteome is less diverse because the heterogeneity in composition was not observed.

**Figure 1:**
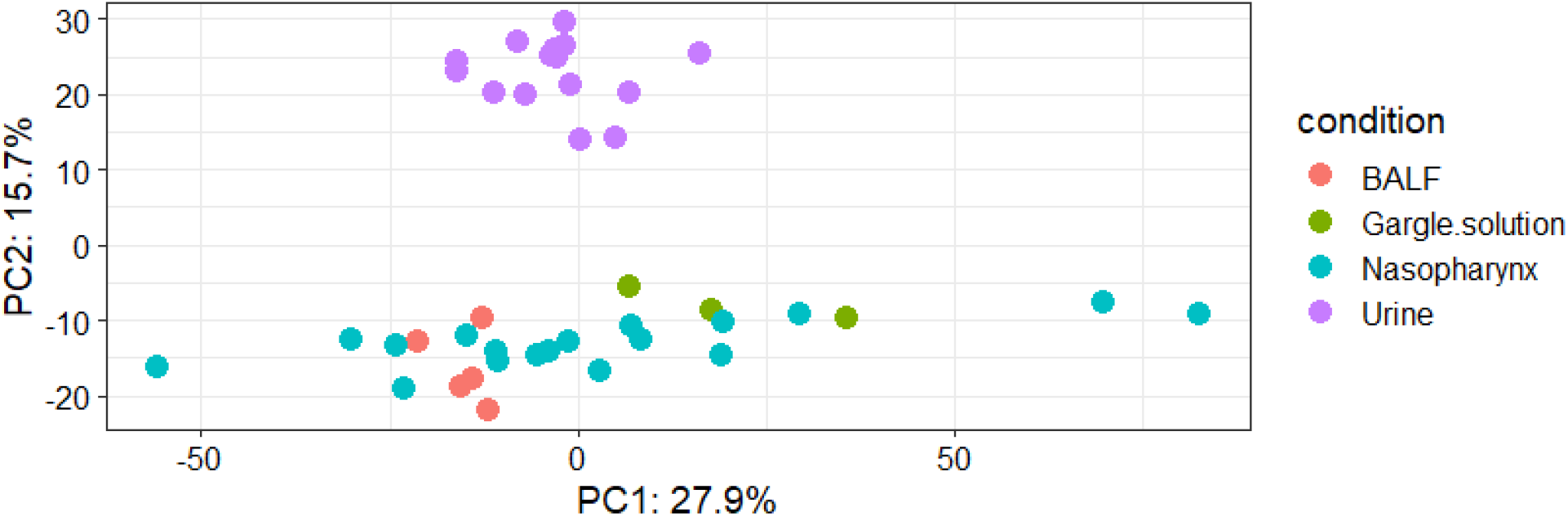
Principal component analysis showing the clustering of proteomes obtained from bronchoalveolar lavage fluid, gargle solution, Nasopharynx, and urine samples

The lack of diversity was also evident in the BALF and indication that there is a coordinated protein secretion in the lungs of the individuals infected with SARS-CoV-2 virus. The gargle showed heterogeneity in proteome composition and indication that the gargle solution has a relative diverse proteome profile in the SARS-CoV-2 seropositive samples. Interestingly, the nasopharyngeal samples demonstrated a high diversity and heterogeneity on proteome composition compared with the data obtained from the other body sites [Figure 1]. Our analysis of proteins obtained from the individuals with confirmed SARS-CoV-2 infections shows that the different body sites respond differently to SARS-CoV-2 antigens.

### Body sites are characterized by different proteins abundance post-SARS-CoV-2 infection

We investigated the protein abundance across the four different samples. Our data demonstrate a clear clustering of individuals into two main clusters [Figure 2]. The difference in the cluster shows that the proteome profiles of the different body sites are the same what differs is their abundances [Figure 2]. Using K-means clustering of proteins, we identified six main protein clusters. Cluster 1 proteins were less abundant in the urine and nasopharynx samples.

**Figure 2:**
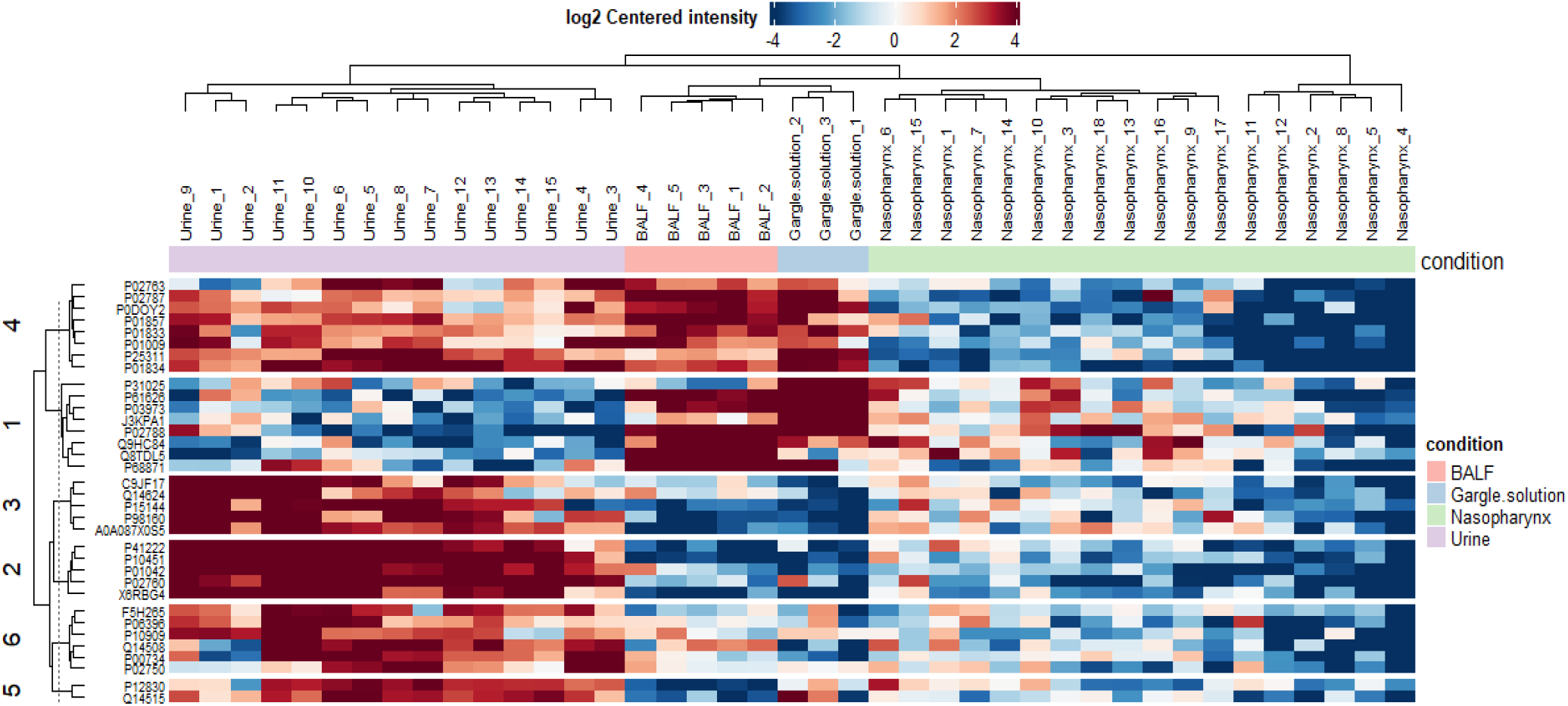
Heatmap showing the protein abundance in urine, bronchoalveolar lavage fluid, gargle solution, and nasopharynx samples we analyzed.

In contrast, the cluster 1 proteins were more abundant in the bronchoalveolar lavage fluid and gargle solution samples [Figure 2]. Proteins in clusters 2,3, 5, and 6 were more abundant in the urine samples. Interestingly, these proteins (in clusters 2,3,4 and 6) were less abundant in the bronchoalveolar lavage fluid, gargle solution, and nasopharynx samples [Figure 2]. Proteins in cluster 4 were more abundant in the urine, bronchoalveolar fluid, and gargle solution, and these proteins were less abundant in the nasopharynx samples [Figure 2].

### A unique set of proteins is dominating human body sites during SARS-CoV-2 infection

The union analysis using <u>Venny</u> 2.1 was conducted to determine the unique and overlapping proteins from the four different body sites we compared [Figure 3]. Urine samples had 257 (33.7%) proteins unique to that body site. We identified 250 (32.8%) proteins uniquely identified in the nasopharynx protein samples. The gargle solution was characterized by a low number of unique identified proteins; 38 (5%) of the identified proteins were unique. The BALF had 73 (9.6%) of the identified proteins following our analysis. Our data shows that the less diverse samples, urine, BALF, and gargle solutions, have a different set of unique proteins in the SARS-CoV-2 infected individuals.

**Figure 3:**
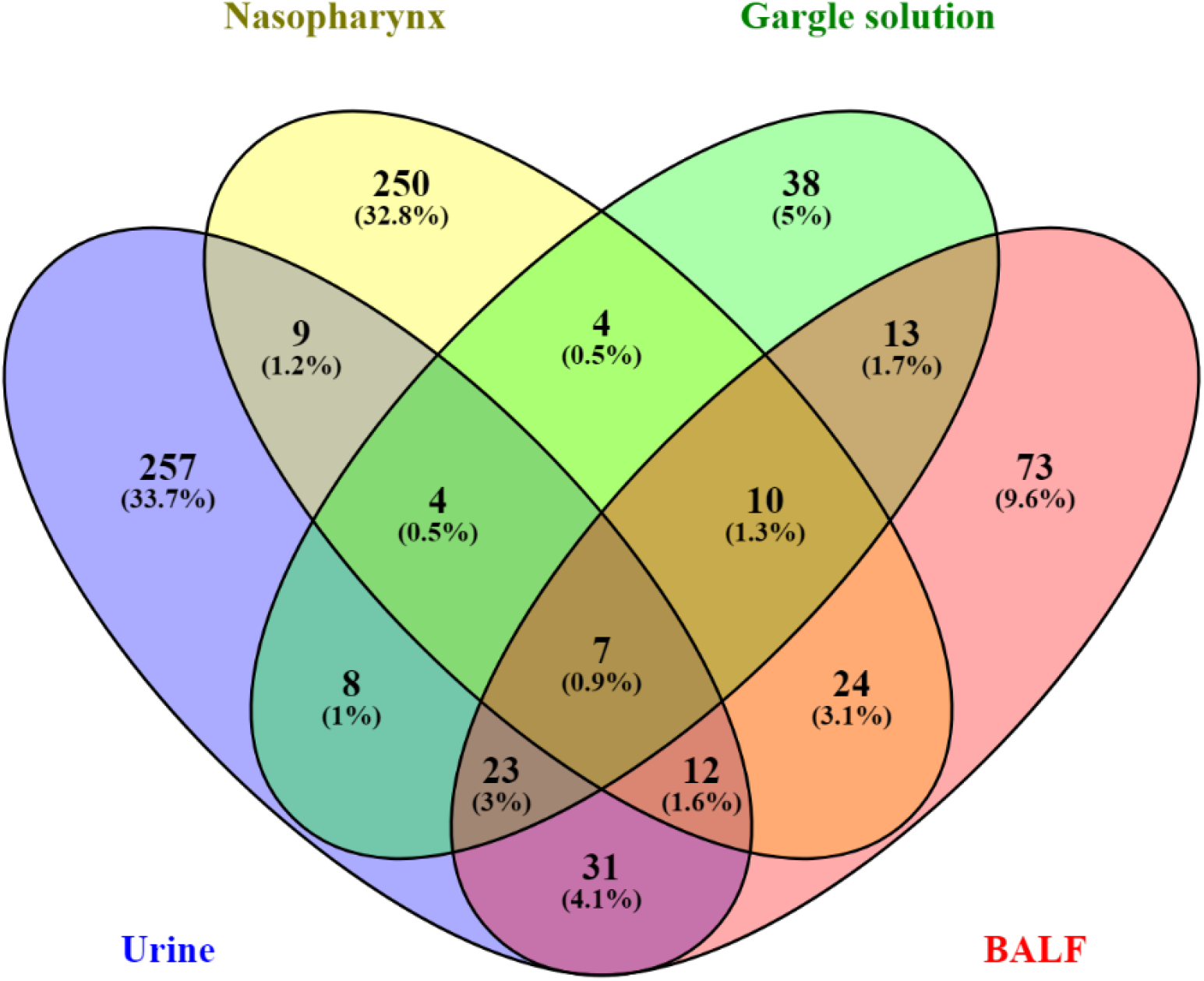
Venn diagram showing the uniques and overlapping proteins identified from urine (blue), Nasopharynx(yellow), Gargle solution (yellow), and BALF (red).

### Different body sites have different sets of regulated proteins post-SARS-CoV-2 infection in the human host

We compared the different body sites to identify the regulated proteins in the different body sites post-SARS-CoV-2 infections in humans [see Figure 4 A-F]. There were 101 upregulated proteins in the gargle solution and 97 downregulated proteins in the BALF when the BALF proteome profile was compared with the gargle solution [Figure 4 A]. Comparing BALF and nasopharynx proteome profiles, 441 proteins were upregulated in the Nasopharynx, and 138 proteins were downregulated in the BALF [Figure 4 B]. There were 331 upregulated proteins in the urine samples compared with the 118 proteins downregulated in the BALF when the BALF proteome was compared with the urine samples [Figure 4 C]. Comparing gargle solution and nasopharynx data, we identified 52 significantly upregulated proteins in the Nasopharynx and 104 proteins being downregulated in the gargle solution [Figure 4 D]. We then compared gargle solution and the urine samples, the two distant body sites, to identify the regulated proteins. Interestingly, 144 and 95 proteins were upregulated in urine and gargle solutions, respectively [Figure 4 E]. Finally, we compared the Nasopharynx and the urine samples, and 166 proteins were upregulated in the urine samples compared with one downregulated protein in the nasopharynx samples [Figure 4 F].

**Figure 4:**
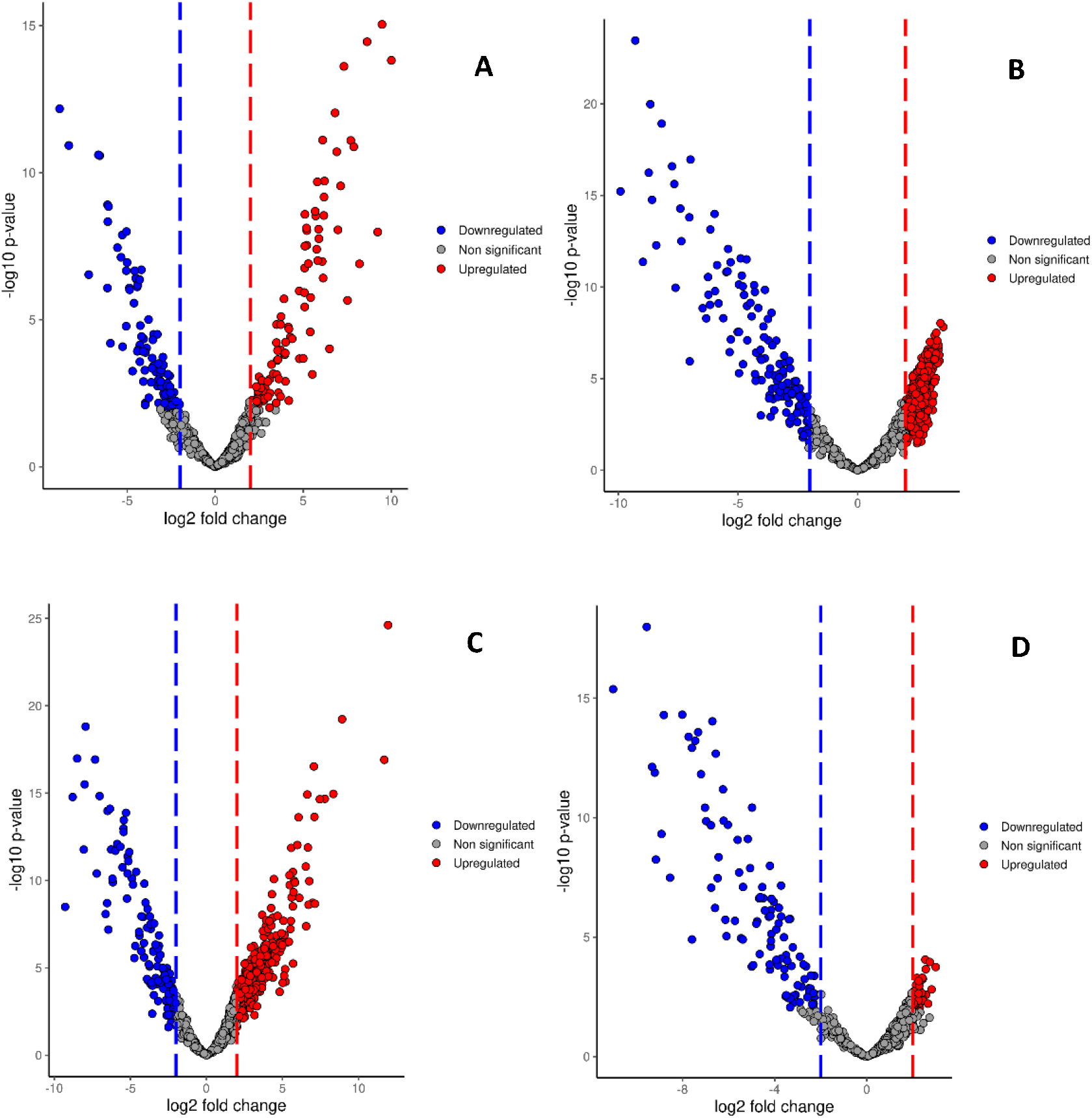
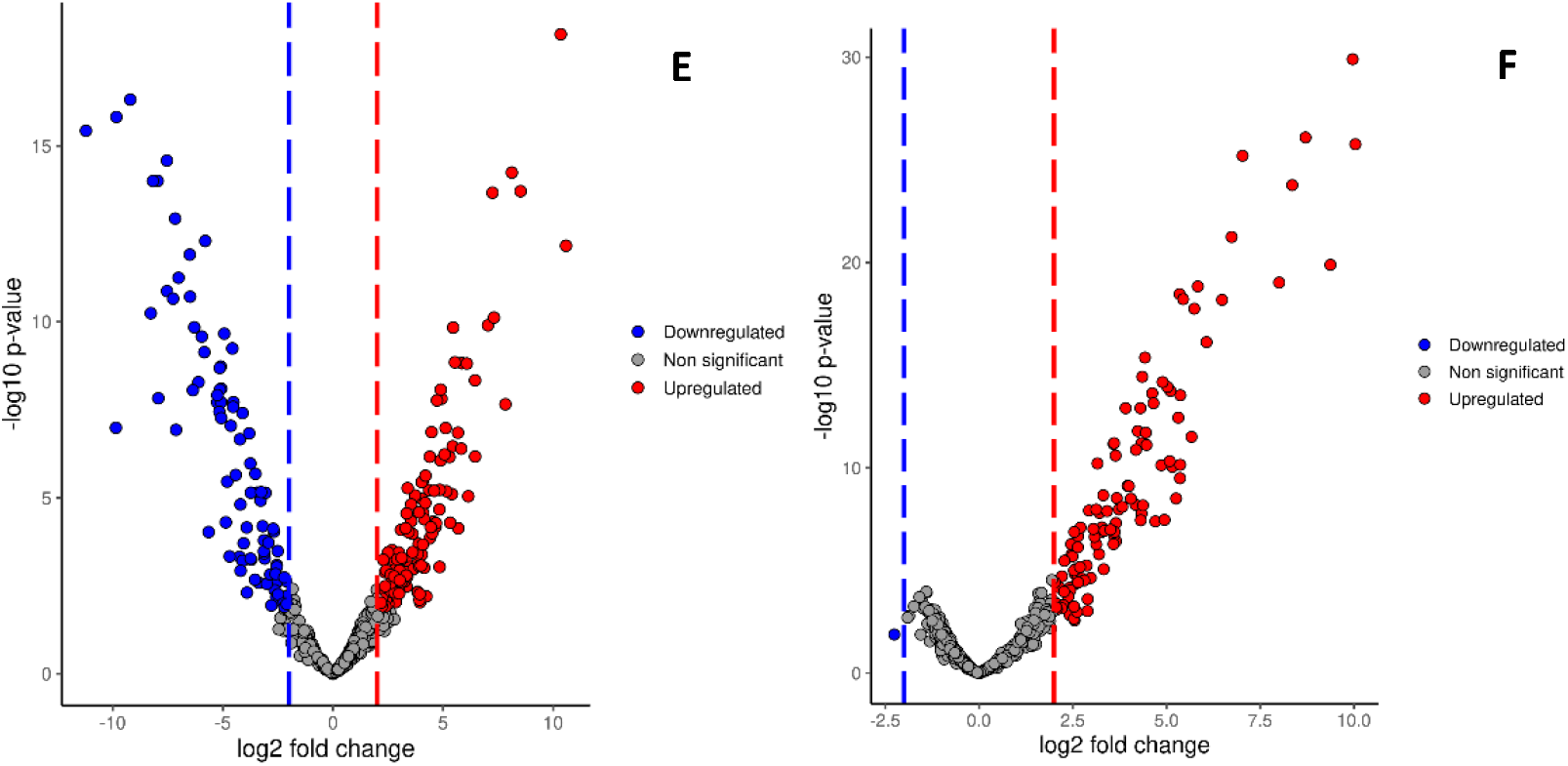
Volcano plots showing regulated proteins in different body sites post-SARS-COV-2 infections in the human host. (A) shows regulated proteins in BALF vs. gargle solution, (B) BALF vs. Nasopharynx, (C) BALF vs. urine, (D) gargle solution vs. Nasopharynx, (E) gargle solution vs. urine, and (F) nasopharynx vs. urine.

## 5. Discussion

Proteomics analysis of SARS-CoV-2 data has been used to identify the potential therapeutic targets in human hosts (19), a practical approach in combating the control and spread of SARS-CoV-2 in our population. The development of testing kits has been made possible due to the use of proteomic data to study and understand SARS-CoV-2 proteomics biomarkers (20,21). It has also been effective in identifying variants with multiple mutations at the immunodominant spike protein, facilitating viral cell entry through the ACE2 receptor (22). This study describes the proteomic profile of the Nasopharynx, gargle solution, urine, and bronchoalveolar lavage fluid obtained from individuals with confirmed SARS-CoV-2 infections (7,8,10,23). The SARS-CoV-2 infection was confirmed by reverse transcriptase-polymerase chain reaction (RT-PCR). PCA reveals a differential diversity of proteins in the investigated body sites. The urine, BALF, and gargle solution proteome profiles demonstrated low diversity, while the nasopharynx proteome data showed a high diversity since they did not cluster together in space. The difference in the proteome diversity can be attributed to the fact that SARS-CoV-2 affects the different body sites differently, as was demonstrated by Feng et al. 2020 (24). The proteomic data obtained from the Nasopharynx demonstrated a high diversity, and this could be explained in part due to heterogeneity of “angiotensin-converting enzyme 2 (ACE2) expression and tissue susceptibility to SARS-CoV-2 infection” (25). Another body of evidence shows that the high diversity of proteome in the nasopharynx samples could be attributed to the impact of the virus on the microbiome (26).

The protein abundance was different in different body sites. Most proteins in clusters 1,3,4,5, and 6 [Figure 2] were more abundant in the urine samples than in the Nasopharynx, gargle solution, and the BALF. The more abundant proteins in the urine samples were also detected in more abundance in the BALF and gargle solution in clusters 1 and 4 [Figure 2]. Chavan et al. 2021 (27) demonstrated that the urine proteome was more differentially expressed in the SARS-CoV-2 cases than the negative control. The difference in abundance and diversity of the proteome profile can be due to the SARS-CoV-2 protein source. In the urine samples, the nucleocapsid protein is the predominant source of protein hence the lack of proteome diversity in this body site (27).

The union analysis reveals a unique set of proteins that characterize human hosts different body sites post-SARS-CoV-2 infections. On the other hand, there were overlaps of the identified proteins from the different body sites with various percentages. Urine samples had the largest number of unique proteins (n=257; 33.7%), followed by the nasopharynx (n=250; 33.8%). The gargle solution had the lowest number of the identified proteins even though the SARS-CoV-2 peptides could still be identified in the gargle solution, making it an important alternative source of samples for SARS-CoV-2 testing. Urine and gargle solution samples can be used to identify and characterize the SARS-CoV-2 virus since they are less invasive and easy to obtain, unlike nasopharyngeal samples.

Urine, gargle solution, BALF, and nasopharyngeal samples demonstrated the difference in the regulated proteins. These differences need to be elucidated, and their clinical relevance needs further investigation. We hypothesize that this significantly regulated protein difference could hamper drug development. The clinical trials should factor the multi-organ comparisons of the proteome profiles of the individuals infected with the SARS-CoV-2 virus in the design and development of anti-SARS-CoV-2 drugs.

In conclusion, we demonstrated that the different body sites have different protein diversity in individuals with confirmed RT-PCR SARS-CoV-2 infection. This study is a proof-of-concept study demonstrating that for the effective design and development of SARS-CoV-2 anti-viral drugs, the protein profiles of the different body sites need to be considered. The finding in this study could have a direct implication on performing population-wide effects of SARS-CoV-2 infections in different body sites.

This study had some limitations. The first limitation is the small sample size, but this did not affect our understanding of the biology under study. We also acknowledge the difference in the sample preparation protocols, which could also be a potential confounder. This is secondary data analysis; we did not have sufficient study participants’ information, and we acknowledge this because it can contribute to the inaccurate interpretation of the results.

## Data Availability

All data produced are available online at https://doi.org/10.6084/m9.figshare.17198384.v2

https://doi.org/10.6084/m9.figshare.17198384.v2

## 6. Abbreviations

ACE2: Angiotensin-Converting Enzyme 2
AKI: Acute Kidney Injury
BALF: Bronchoalveolar lavage fluid
FDR: False Discovery Rate
IPX: Integrated Proteome resources
LFQ: Label-Free Quantitation
PCA: Principal Component Analysis
PRIDE: Protein Identification Database
PSM: Peptide Spectral Match
SARS-CoV-2: Severe Acute Respiratory Syndrome Coronavirus 2.

## Notes

### Competing Interest Statement

The authors have declared no competing interest.

### Funding Statement

This study did not receive any funding

### Author Declarations

This study involves only openly available human data, which can be obtained from: https://www.ebi.ac.uk/pride/ with the following accession numbers: PXD019423, PXD018970, PXD022085, and PXD022889

